# Neck Vibration-Evoked Nystagmus in Vestibular Migraine: Mechanistic Insights into Role of Proprioception

**DOI:** 10.64898/2026.03.10.26348105

**Authors:** Francisco Zuma e Maia, Bernardo Faria Ramos, Jorge Otero Millan, Marcio Cavalcante Salmito, Renato Cal, Sofiene ben Rhouma, Pierre Miniconi, Aasef G Shaikh

## Abstract

**Introduction:** Vestibular migraine is a major cause of recurrent vertigo, yet its mechanisms and diagnostic markers remain limited. Abnormal vestibular–cervical integration and convergence insufficiency, reflected by impaired near point convergence (NPC),suggest multisensory dysfunction. This study tested whether cervical proprioceptive perturbation provokes vertigo and nystagmus in vestibular migraine and evaluated NPC as a predictor of these responses.

**Methods:** Fifty-one vestibular migraine patients and 12 controls underwent interictal vestibular testing. Peripheral function was assessed with vHIT. Participants received randomized 100-Hz cervical (proprioceptive) and mastoid (vestibular) vibration without visual fixation, with eye movements recorded via video Frenzel goggles and NPC measured using standard methods. Analyses included McNemar’s, Wilcoxon signed-rank, Mann–Whitney U, correlations, and multivariable logistic regression.

**Results:** Neck vibration provoked vertigo in all vestibular migraine patients and none of the controls, producing nystagmus in 76.5%. Horizontal, ipsiversive nystagmus predominated, while less frequent vertical responses showed higher velocities. Mastoid vibration elicited no nystagmus. NPC was the only independent predictor of nystagmus and correlated with slow-phase velocity and bilateral responses. Age correlated with drift velocity, whereas vestibulo-ocular reflex gain showed no association.

**Discussion:** Neck vibration elicits vertigo and nystagmus in vestibular migraine, providing the first objective physiological marker. NPC predicts and correlates with nystagmus severity, highlighting its value as a surrogate of multisensory dysfunction. Together, these findings implicate abnormal cervical–vestibular integration and position NPC and neck-vibration testing as practical tools for diagnosis and phenotyping.

**Key points:**

- Vestibular migraine affects ∼3% of population yet remains highly controversial.
- Objective measures reveal reproducible vertigo and nystagmus in vestibular migraine.
- Impaired convergence strongly predicts vibration-induced nystagmus in VM patients.
- Findings support sensory mismatch model linking cervical proprioception to vertigo.

## Introduction

Aretaeus of Cappadocia first noted an association between migraine and vertigo in 131 BC, but systematic studies began in the late 19th century^1^; since 1990 publications on vertigo and headache have roughly doubled every five years. The variety of terms—vestibular migraine (VM)^2^, migranous vertigo, migraine-associated vertigo, vertiginous migraine, and others reflected historical controversy, not only semantic disagreement but a deeper debate: is this a single disorder with a recognizable signature or a spectrum of overlapping conditions that simply co-occur?

Vertigo affects about 7.4% of the general population^3^, while migraine affects roughly 12–16%^3, 4^. If unrelated, their co-occurrence would be approximately 1%, yet the observed rate is 3.2%, about three times higher, supporting a non-random association^4–6^. These epidemiological findings that co-occurrence exceeds expected prevalence sharpened focus on the migraine–vestibular link and fueled the view that migraine-related vestibular symptoms may be a leading cause of vertigo^3^.

VM was formally listed as a distinct entity in the appendix of the International Classification of Headache Disorders 3-beta (ICHD-3 beta, A1.6.5), following a consensus by the International Headache Society and the Bárány Society that brought neurologists and otorhinolaryngologists together to set diagnostic criteria^7^. The defining criteria were vestibular symptoms lasting between 5 minutes to 72 hours, occurring during at least 50% of migraine episodes, and accompanied by migraine-associated features such as headache, photophobia, phonophobia, or aura^8^. Beyond these core criteria, VM presents with higher rates of occipital headache and cervical muscle tension than migraineurs without vestibular involvement^9^. Convergence insufficiency is frequently observed, leading to impaired depth perception and dizziness^10^. Nystagmus is not uncommon during acute attacks and has also been reported between attacks^11–14^. Neck pain from cervical muscle spasm affects roughly 37–46% of patients, further compounding imbalance. Such breadth of symptomatic features has generated a concern of limited specificity of VM as a diagnosis in the absence of objective physiological markers to separate VM from other vestibular disorders and migraine phenotypes^15^.

Not just the physiological markers, but the underlying mechanisms also remain elusive. The constellation of symptoms, ranging from vertigo and imbalance to convergence insufficiency and neck pain and spasms, suggests that VM is not merely a disorder of isolated vestibular dysfunction but rather a condition involving multisensory integration. This concept deserves further investigations. The clinical observations raise critical questions about how vestibular, visual, and neck proprioceptive signals interact in VM and whether abnormal processing within these systems contributes to generation of vestibular symptoms^16^. Addressing these gaps in pathophysiology and finding an objective physiological marker for VM motivated our investigation into the role of abnormal cervical proprioception in the pathophysiology of VM.

We hypothesized that vertigo in VM arises from abnormal processing of neck proprioceptive input. This aberrant sensory integration may persist in between migraine attacks but becomes markedly exaggerated during acute migraine episodes, producing noxious symptoms of unsteadiness and vertigo. In severe cases, this dysfunction may even manifest as nystagmus.

Our hypothesis made a clear, testable prediction: abnormal proprioceptive–vestibular interactions in VM, manifesting as acute vertigo or nystagmus, can be provoked in a controlled clinical setting by externally altering cervical proprioception. We used neck muscle vibration, a well-characterized method that perturbs cervical proprioceptive input, to test this prediction and anticipated that it would trigger on-demand exacerbations of vertigo and, in some cases, nystagmus in individuals with VM. To evaluate this, we enrolled 51 patients meeting VM diagnostic criteria and 12 healthy controls and applied the cervical vibration protocol while recording vestibular and oculomotor responses. The intervention selectively and reproducibly elicited vertigo and nystagmus in VM patients but not in controls, providing the first objective physiological marker that distinguishes VM, enables on-demand phenotyping, and offers a practical tool for diagnosis and mechanistic study.

## Materials and Methods

We studied 51 participants (40 women) diagnosed with vestibular migraine (VM) and 12 healthy controls (8 women). The average age of VM participants was 51.9 ± 17.5 years; while it was 36.8±17.6 year for the healthy controls. VM was diagnosed according to the Bárány Society and International Headache Society criteria, as noted above ^8^. The study was approved by the institutional ethics committee; all participants provided written informed consent prior to enrollment. Experiments were conducted during the interictal phase, when participants were free of acute migraine symptoms. Each participant completed a single experimental session consisting of three phases:

1. The integrity of the peripheral vestibular system was evaluated using the video head impulse test (Synapsys).
2. Mastoid vibration at 100 Hz was applied using the FRAMIRAL system (George Dumas Test, Grenoble, France). Stimulation was delivered sequentially to the left and right mastoid processes, just above the insertion of the sternocleidomastoid muscle, in randomized order while participants were seated without visual fixation.
3. Cervical vibration at 100 Hz was applied using the FRAMIRAL system (Pierre Miniconi Test, Carpentras, France). Stimulation was delivered sequentially to the left and right sides of the neck (5 cm below the inion, 2 cm lateral), in randomized order while seated without visual fixation.

Eye movements were recorded throughout using video Frenzel goggles^17^. Analyses focused on presence of nystagmus, its direction, and slow-phase velocity. Near point convergence (NPC) was measured while participants sat upright and fixated on the target held ∼50 cm in the midsagittal plane. The examiner advanced the target smoothly toward the nose until NPC was reached, then moved it back until binocular fixation resumed. The test was performed under good illumination to detect eye movement changes. Statistical methods included binomial confidence intervals, McNemar’s test, Wilcoxon signed rank, Mann–Whitney U, Spearman/Pearson correlations, and logistic regression with VOR gain, NPC, age, and sex as predictors. Model performance was evaluated by AUC and VIF; all tests were two sided at α = 0.05.

## Results

All 63 participants—51 with vestibular migraine (VM) and 12 healthy controls—successfully completed the full study protocol. Figure 1A shows healthy control during mastoid vibration (top panel, Figure 1A). Here the mastoid, which modulates inner ear end-organ output, does not disrupt gaze stability. The bottom panel of Figure 1A presents gaze-holding during neck vibration applied at standardized anatomical landmarks (see Methods). Again, no gaze instability is observed. VOR gain in this participant was normal. Figures 1B depicts responses from a representative VM patient. As in controls, mastoid vibration (top panel Figure 1B) was associated with stable gaze. However, left-sided neck vibration (bottom panel Figure 1B) elicits downward drifts, i.e., upbeat nystagmus. The patient reported intense vertigo simultaneously during neck vibration. The VOR during vHIT was normal.

**Figure 1:**
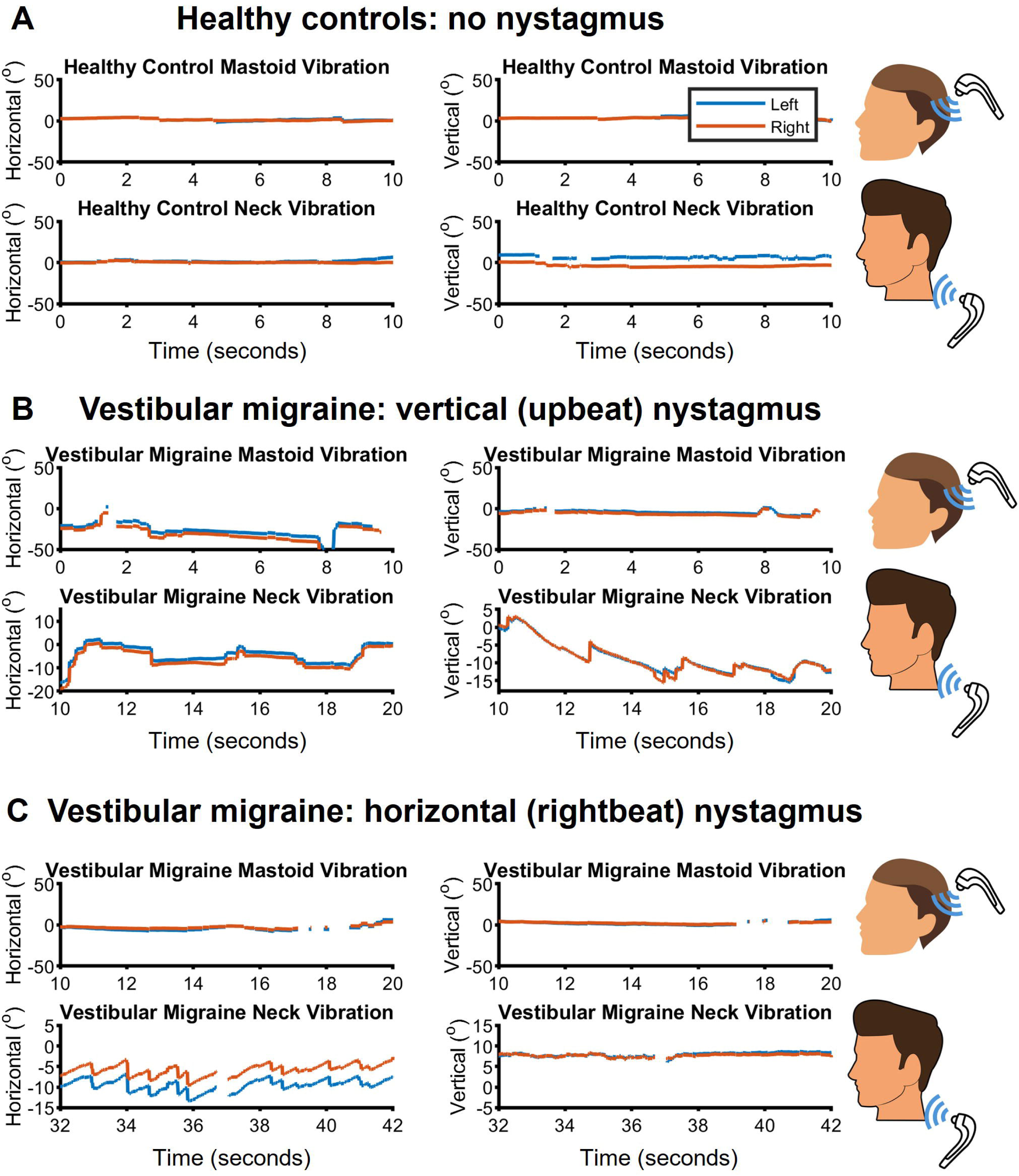
Example of gaze holding during mastoid vibration and neck vibration in healthy participant (A), and two examples of vestibular migraine patients (B: example 1; and C: example 2). In each plot y-axis depicts angular eye position (vertical or horizontal) while x-axis illustrates corresponding time. Arrows in panels B and C depicts nystagmus.

Figures 1C show data from another VM patient. Mastoid vibration did not affect gaze stability (top panel Figure 1C), and vHIT was normal. However, right-sided neck vibration (bottom panel Figure 1C) induced left-beat nystagmus. The patient reported sensation of vertigo simultaneously during neck vibration.

These cases demonstrate that while mastoid vibration does not affect gaze stability in either healthy controls or VM patients, neck vibration can elicit distinct patterns of nystagmus and vertiginous sensations. To further evaluate the consistency and specificity of these findings, we analyzed responses across the full cohort of 51 VM patients. We specifically asked:

1. What is the prevalence of vertigo and nystagmus with neck vibration.
2. Which parameters predict presence of nystagmus.
3. Which parameters correlate with the intensity of the nystagmus.

### Perceived Vertigo During Neck Vibration

Neck vibration consistently evoked a subjective sensation of vertigo in all VM patients. The experience was described as a spinning sensation or profound unsteadiness.

### Prevalence of Nystagmus During Neck Vibration

Nystagmus was observed in 39 of 51 (76.5%) VM participants during neck vibration. Horizontal responses were significantly more prevalent than vertical responses for each side (Right: 52.9% vs 17.6%; Left: 54.9% vs 19.6%) (Figure 2A). McNemar’s exact test confirmed this difference (Right: 20 horizontal-only vs 2 vertical-only, p = 1.2 × 10⁻⁴; Left: 19 vs 1, p = 4.0 × 10⁻⁵), indicating a horizontal bias.

**Figure 2:**
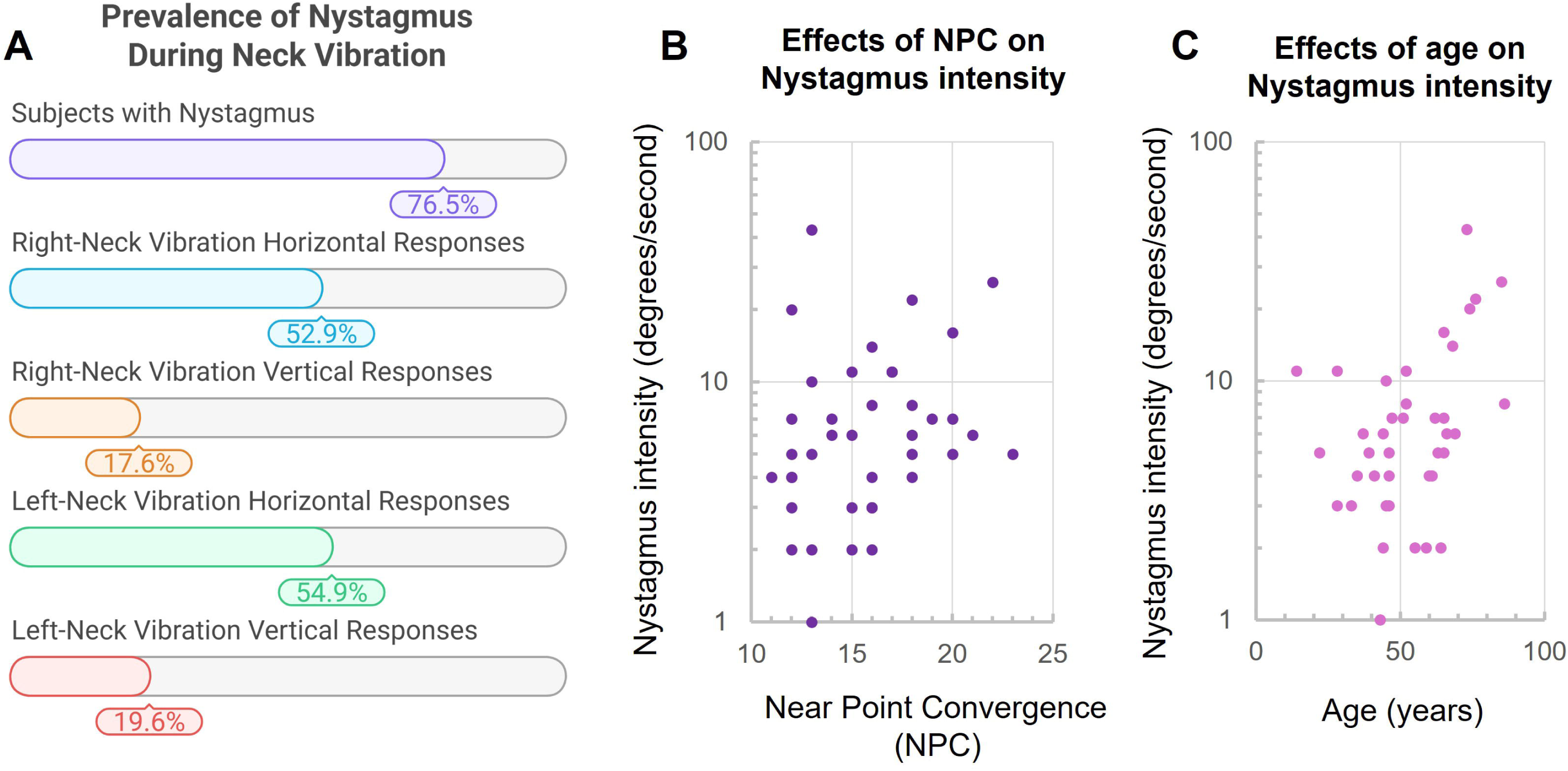
(A) Summary of incidence of vibration induced nystagmus and its laterality. Correlation analysis depicting (B) relationship between nystagmus intensity and near point convergence (NPC); and (C) nystagmus intensity and age of the participant. In panels B,C each symbol depicts one participant, it is to be noted that the correlation analysis is only performed in cases where nystagmus was present.

During right-neck vibration, ipsiversive rightward drift was most frequent (37.3% of subjects; mean velocity 3.74°/s, median 4°/s), followed by contraversive leftward drift (15.7%; mean 4.00°/s). Vertical responses were rare (Up: 9.8%; Down: 7.8%) but upward drift had the highest velocities (mean 8.00°/s; max 21°/s). During left-neck vibration, ipsiversive leftward drift predominated (39.2%; mean 3.65°/s), contraversive rightward drift occurred in 15.7% (mean 4.88°/s), and upward drift reached the highest velocity (22°/s). Ipsiversive responses were more common than contraversive for both sides, but McNemar’s test showed significance only for left-neck vibration (Right: 19 vs 8, p = 0.052; Left: 20 vs 8, p = 0.036). Paired Wilcoxon tests comparing drift velocity magnitudes revealed a trend toward ipsiversive dominance for right-neck vibration (n = 27, p = 0.071, rank-biserial r = 0.40) and a significant effect for left-neck vibration (n = 28, p = 0.019, r = 0.50).

Within-subject comparisons of maximum horizontal vs vertical drift velocity showed horizontal dominance for most subjects (Left: n = 29, p = 0.0021, r = 0.65; Right: n = 24, p = 0.0028, r = 0.69). Although horizontal maxima were typically greater, the largest absolute velocities occurred in vertical-upward drifts (21–22°/s), indicating a heavy-tailed vertical distribution.

### Predictive Modeling

In this cohort (N = 51; 39/51 = 76.5% with nystagmus), a multivariable logistic regression including age, near point of convergence (NPC), sex (M/F), right VOR gain, and left VOR gain identified NPC as the only significant independent predictor of nystagmus (OR per +1 unit = 1.52, 95% CI 1.04–2.21, p = 0.031)(Table 1). Age (OR 1.03, 95% CI 0.98–1.07, p = 0.28), and male sex (OR 2.67, 95% CI 0.33–21.87, p = 0.36) were not significant, and the model showed moderate discrimination (AUC 0.75). Univariately, NPC showed a trend (OR per +1 = 1.26, 95% CI 0.97–1.63, p = 0.085; AUC 0.67).

**Table 1.** Statistical summary of multilinear regression analysis depicting near point convergence (NPC) as a strong predictor for the presence of nystagmus.

| Predictor | Odds Ratio (per unit) | 95% confidence interval | p-value |
| --- | --- | --- | --- |
| Near point convergence (NPC) | 1.52 | 1.04-2.21 | 0.031* |
| Age | 1.03 | 0.98-1.07 | 0.279 |
| Male Vs. Female | 2.67 | 0.3-21.9 | 0.359 |
| Right VOR Gain | $4.34 \times 10^4$ | $2.95 \times 10^{-5}$-$6.39 \times 10^{13}$ | 0.321 |
| Left VOR Gain | 0.2 | $1.11 \times 10^{-10}$-$3.46 \times 10^8$ | 0.881 |

### Correlation of Nystagmus Intensity with NPC

The subsequent analysis examined, when present, how nystagmus slow-phase velocity correlates with the severity of impaired NPC (Figure 2B), how often nystagmus occurs on both sides versus only one side, and whether vertical components (upbeat/downbeat) appear bilaterally or unilaterally. Drift velocity, the intensity metric, positively correlated with NPC during right-side neck vibration when summarized as the total drift across directions (Spearman ρ = 0.433, p = 0.001; Pearson r = 0.269, p = 0.056) and as the maximum leftward drift (ρ = 0.391, p = 0.005; Pearson r = 0.182, p = 0.201). NPC also correlated with the overall total drift pooled across both sides (ρ = 0.359, p = 0.01; Pearson r = 0.215, p = 0.130). Associations of drift velocity during left-neck vibration were smaller and not significant (Left-total: ρ = 0.149, p = 0.298; Left-max: ρ = 0.102, p = 0.474); while the overall maximum drift showed a weaker, non-significant trend (ρ = 0.244, p = 0.084).

Regarding laterality of occurrence, nystagmus was present bilaterally (both right- and left-neck vibration) in 19/51 (37.3%), unilaterally (only one side) in 20/51 (39.2%), and absent in 12/51 (23.5%). NPC differed across these patterns; those with bilateral responses had higher NPC (mean ± SD 16.68 ± 3.15; median 17.0; n = 19) than those with no nystagmus (13.92 ± 2.47; median 13.5; n = 12; Mann–Whitney U = 172.0, p = 0.019), and showed a borderline elevation versus unilateral responders (14.80 ± 2.97; median 15.0; n = 20; U = 259.5, p = 0.0508). Taken together, higher values of NPC correlate with greater intensity of neck vibration-induced nystagmus.

### Correlation with Age and Drift Amplitude

Analysis revealed that age was positively associated with overall drift magnitude, particularly when expressed as the maximum velocity observed across all directions and both sides (Figure 2C). Spearman correlation between age and overall maximum drift was ρ = 0.395 (p = 0.004; BH-adjusted q = 0.064), corroborated by Pearson’s r = 0.446 (p = 0.001), indicating that older subjects tended to exhibit higher peak drift velocities. Overall total drift also showed a weaker but significant unadjusted association (ρ = 0.296, p = 0.035), though this did not survive multiple-comparison correction. Side-specific analyses revealed similar trends for right- and left-side maxima (ρ ≈ 0.24–0.26), but these were not statistically robust after adjustment. Direction-specific correlations highlighted a selective effect for left-side downward drift, which increased with age (ρ = 0.372, p = 0.007, q = 0.064), whereas other directions—including contraversive and ipsiversive horizontal drifts—showed no significant age dependence. Directionality metrics, including contraversive–ipsiversive differences and horizontal–vertical bias, were unrelated to age (all p > 0.40), and Mann–Whitney comparisons confirmed no age difference between subjects with contraversive dominance versus those without. Collectively, these findings suggest that older individuals are more likely to have greater drift velocities overall, with a possible contribution from vertical-downward components on the left, while the directional preference of nystagmus remains largely age-independent.

To summarize, transient perturbation of neck proprioception by means of neck vibration instantaneously triggers acute vertigo and nystagmus. Convergence impairment measured with NPC is a strong predictor of neck-vibration induced nystagmus. The intensity of nystagmus correlates with severity of NPC and the patients’ age. The results provide first objective marker of VM, while depicting the role of aberrant multisensory integration of vestibular and proprioceptive input as a cause of VM.

## Discussion

The overarching goal of our investigation is to highlight a novel pathophysiological feature and a putative diagnostic marker of VM. The deficit highlights, on demand, transient provocation of intense percept of vertigo, frequently accompanied by nystagmus; both induced by neck vibration that is thought to transiently modulate neck proprioception. The nystagmus observed was present in horizontal (right or left) or vertical (up or down) planes. Horizontal nystagmus was often associated with lateralized neck vibration, appearing only during stimulation of the left or right side, and consistently drifting toward the vibrated side. Vertical nystagmus, though less common, occurred with both unilateral and bilateral neck vibration. Peripheral vestibular system was normal in VM as well as healthy participants. Mastoid vibration, which mechanically stimulates the labyrinth in an asymmetric fashion did not trigger nystagmus in VM, as expected from healthy participants. These features are consistent with a healthy vestibular system^18, 19^. However, observations under neck vibration prompted further exploration of the physiological basis underlying exaggerated responses to neck vibration.

The results aligned with previous investigations that examined vestibular evoked myogenic potentials (VEMPs) and vestibulo-collic reflexes (VCR). Cervical VEMP amplitudes are higher in VM but not significantly different from migraine without vestibular symptoms^19^. Ocular VEMPs, however, were consistently prolonged in VM, implicating utriculo-ocular pathway involvement^20, 21^. Further, VCR amplitudes are significantly reduced in interictal migraine patients compared to controls, while migraineurs exhibit potentiation rather than habituation with repeated stimuli^22, 23^. These findings point to abnormal central processing of repeated vestibular stimuli, a hallmark of migraine pathophysiology, rather than peripheral hyperexcitability. Foam posturography studies corroborate this, showing significantly greater postural instability in VM patients compared to healthy controls, reinforcing the role of proprioceptive dysfunction in VM^24^.

Neck vibration (60–100 Hz) stimulates cervical muscles. In healthy individuals, it may cause perceptual illusions like room tilt but rarely instability^25, 26^. In VM, responses are amplified and prolonged, with instability persisting even between attacks, suggesting chronic proprioceptive deficits. We propose abnormal integration of cervical proprioceptive and vestibular signals underlies these findings; disrupting balance and reflex control. Left-sided vibration induced right-beating horizontal and occasional vertical nystagmus, while right-sided vibration produced the opposite. This pattern is analogous to compensated unilateral vestibular hypofunction, where non-physiological maneuvers such as skull vibration can re-evoke latent asymmetry, offering mechanistic insight into abnormal neck proprioception in VM. The concept is further elaborated below.

Vestibular afferents converge with neck proprioceptive inputs at the vestibular nuclei, generating integrated signals essential for motion perception, gaze stabilization, and the VOR^27, 28^. In acute vestibular hypofunction, nystagmus drifts toward the lesioned ear and compensates over time, but non-physiological stimuli such as skull vibration can re-evoke latent asymmetry, producing nystagmus similar to the acute phase^18, 29, 30^. We speculate that VM reflects a comparable mechanism, with asymmetric neck proprioception creating skewed vestibular-proprioceptive output despite normal vestibular tone. Chronic adaptation may mask this imbalance, but vibration or migraine-related muscle stiffness can acutely reinstate asymmetry, triggering vertigo or nystagmus. Alternatively, excessive proprioceptive feedback may suppress vestibular output, causing vertigo acutely and persistent unsteadiness chronically—both characteristic of VM. Furthermore, central sensitization in VM may amplify proprioceptive signals, explaining why neck vibration can provoke symptoms resembling spontaneous VM attacks.

Impaired integration of neck proprioceptive input may account for the frequent co-occurrence of neck pain, cervical stiffness, and disequilibrium in migraineurs, even during interictal periods. Altered axial proprioception likely contributes to heightened motion sensitivity and postural instability, both hallmark features of VM. Collectively, these findings, reinforced by our results, suggest that VM involves not only aberrant central processing of vestibular stimuli but also dysfunctional integration of cervical and axial proprioception into postural and reflexive motor control.

Our study highlights the critical intersection between cervical proprioception and vestibular processing in VM, pointing to maladaptive sensory integration that amplifies conflicting signals from the neck and inner ear. These observations strengthen the sensory mismatch model of VM, where cervical dysfunction (whether primary or secondary) exacerbates vestibulo-thalamo-cortical disturbances. Clinically, neck vibration testing emerges as a promising biomarker for diagnosis and subtyping of VM, particularly in patients without classic migraine headaches. These findings also provide mechanistic underpinning further advancing the treatment of VM symptomatic relief. As cervical-vestibular integration becomes increasingly recognized in VM pathophysiology, the “vestibular chameleon” may finally reveal its distinctive patterns.

## Data Availability

All data produced in the present study are available upon reasonable request to the authors

## Conflict of Interests

A. Shaikh is Co-Chief Editor of Dystonia – The Official Journal of Dystonia Medical Research Foundation, Associate Editor of The Cerebellum, and Associate Editor of Neurological Sciences. Shaikh serves as consultant on speaker bureau for Merz Pharmaceuticals. Remaining authors have no conflict of interest to declare.

## Acknowledgements

A.Shaikh was supported by grants from US Department of Veterans Affairs (I01CX002086), Penni and Stephen Weinberg Chair in Brain Health.

## Notes

### Competing Interest Statement

The authors have declared no competing interest.

### Funding Statement

This study did not receive any funding

### Author Declarations

Ethics committee of Federal University Espirito Santo gave ethical approval for this work. Participants provided written informed consent prior to enrollment.

## References

1. Cal R, Bahmad F, Jr. Migraine associated with auditory-vestibular dysfunction. Braz J Otorhinolaryngol. 2008;74:606–612.

2. Dieterich M, Brandt T. Episodic vertigo related to migraine (90 cases): vestibular migraine? J Neurol. 1999;246:883–892.

3. Vukovic V, Plavec D, Galinovic I, Lovrencic-Huzjan A, Budisic M, Demarin V. Prevalence of vertigo, dizziness, and migrainous vertigo in patients with migraine. Headache. 2007;47:1427–1435.

4. Swaminathan A, Smith JH. Migraine and vertigo. Curr Neurol Neurosci Rep. 2015;15:515.

5. Neuhauser HK, von Brevern M, Radtke A, et al. Epidemiology of vestibular vertigo: a neurotologic survey of the general population. Neurology. 2005;65:898–904.

6. Lempert T, Neuhauser H. Epidemiology of vertigo, migraine and vestibular migraine. J Neurol. 2009;256:333–338.

7. Headache Classification Committee of the International Headache S. The International Classification of Headache Disorders, 3rd edition (beta version). Cephalalgia. 2013;33:629–808.

8. Lempert T, Olesen J, Furman J, et al. Vestibular migraine: diagnostic criteria. J Vestib Res. 2012;22:167–172.

9. Wattiez AS, O’Shea SA, Ten Eyck P, et al. Patients With Vestibular Migraine are More Likely to Have Occipital Headaches than those With Migraine Without Vestibular Symptoms. Headache. 2020;60:1581–1591.

10. Maia F, Ramos BF, Bittar RSM, Cal RVR, Luis LA, Albernaz PLM. The Near Point of Convergence in Patients with Vestibular Migraine. Int Arch Otorhinolaryngol. 2025;29:1–4.

11. El-Badry MM, Samy H, Kabel AM, Rafat FM, Sanyelbhaa H. Clinical criteria of positional vertical nystagmus in vestibular migraine. Acta Otolaryngol. 2017;137:720–722.

12. Oh SY, Seo MW, Kim YH, Choi KD, Kim DS, Shin BS. Gaze-evoked and rebound nystagmus in a case of migrainous vertigo. J Neuroophthalmol. 2009;29:26–28.

13. Polensek SH, Tusa RJ. Nystagmus during attacks of vestibular migraine: an aid in diagnosis. Audiol Neurootol. 2010;15:241–246.

14. Gufoni M, Casani AP. “The Pupillary (Hippus) Nystagmus”: A Possible Clinical Hallmark to Support the Diagnosis of Vestibular Migraine. J Clin Med. 2023;12.

15. Stolte B, Holle D, Naegel S, Diener HC, Obermann M. Vestibular migraine. Cephalalgia. 2015;35:262–270.

16. Li Y, Yang L, Dai C, Peng B. Proprioceptive Cervicogenic Dizziness: A Narrative Review of Pathogenesis, Diagnosis, and Treatment. J Clin Med. 2022;11.

17. Strupp M, Arbusow V, Dieterich M, Sautier W, Brandt T. Perceptual and oculomotor eeects of neck muscle vibration in vestibular neuritis. Ipsilateral somatosensory substitution of vestibular function. Brain. 1998;121 ( Pt 4):677–685.

18. Teggi R, Gatti O, Familiari M, Cangiano I, Bussi M. Skull Vibration-Induced Nystagmus Test (SVINT) in Vestibular Migraine and Meniere’s Disease. Audiol Res. 2021;11:603–608.

19. Salmito MC, Gananca FF. Video head impulse test in vestibular migraine. Braz J Otorhinolaryngol. 2021;87:671–677.

20. Kim CH, Jang MU, Choi HC, Sohn JH. Subclinical vestibular dysfunction in migraine patients: a preliminary study of ocular and rectified cervical vestibular evoked myogenic potentials. J Headache Pain. 2015;16:93.

21. Fujimoto C, Kamogashira T, Takenouchi S, et al. Utriculo-ocular pathway dysfunction is more frequent in vestibular migraine than probable vestibular migraine. J Neurol. 2020;267:2340–2346.

22. Allena M, Magis D, De Pasqua V, Schoenen J, Bisdore AR. The vestibulo-collic reflex is abnormal in migraine. Cephalalgia. 2007;27:1150–1155.

23. Roceanu A, Allena M, De Pasqua V, Bisdore A, Schoenen J. Abnormalities of the vestibulo-collic reflex are similar in migraineurs with and without vertigo. Cephalalgia. 2008;28:988–990.

24. Fujimoto C, Koyama M, Kawahara T, et al. Postural stability in patients with vestibular migraine and probable vestibular migraine in the absence of acute vestibular symptoms. Am J Otolaryngol. 2025;46:104551.

25. Taylor JL, McCloskey DI. Illusions of head and visual target displacement induced by vibration of neck muscles. Brain. 1991;114 ( Pt 2):755–759.

26. McKenna GJ, Peng GC, Zee DS. Neck muscle vibration alters visually perceived roll in normals. J Assoc Res Otolaryngol. 2004;5:25–31.

27. Sylvestre PA, Choi JT, Cullen KE. Discharge dynamics of oculomotor neural integrator neurons during conjugate and disjunctive saccades and fixation. J Neurophysiol. 2003;90:739–754.

28. Shaikh AG, Meng H, Angelaki DE. Multiple reference frames for motion in the primate cerebellum. J Neurosci. 2004;24:4491–4497.

29. Waissbluth S, Sepulveda V. The Skull Vibration-induced Nystagmus Test (SVINT) for Vestibular Disorders: A Systematic Review. Otol Neurotol. 2021;42:646–658.

30. Curthoys IS, Zee DS, Dumas G, Pastras CJ, Dlugaiczyk J. Skull vibration induced nystagmus, velocity storage and self-stability. Front Neurol. 2025;16:1533842.

